# *Mycobacterium leprae* infection, Hansen’s disease, and helminth infections: A cross-sectional study in southeastern Brazil

**DOI:** 10.1101/2025.06.26.25330341

**Authors:** Audra Bass, Heloine Leite, Lorena B. P. Oliveira, Emma Nedell, Pedro Marçal, Julie A. Clennon, Jeffrey M. Collins, Thomas R. Ziegler, Erica Magueta Silva, Maisa P. Vieira, Marcos D. S. Pinheiro, Alexandre C. Branco, José A. Ferreira, Lance Waller, Lucia A. O. Fraga, Jessica K. Fairley

## Abstract

**Background:** Prior studies have demonstrated associations between helminths and mycobacterial infections, suggesting a can alter the susceptibility do mycobacterial infection or disease. Our goal was to assess the association of *Mycobacterium leprae* infection with parasitic infections in a highly endemic area for Hansen’s disease in Minas Gerais, Brazil.

**Methods:** Adults and children ages 3 years and older were enrolled from communities in Governador Valadares, Minas Gerais, Brazil, and nearby municipalities. Questionnaires on demographics and infection history were administered. Serological reactivity against *M. leprae* and parasitic infections was assessed by multiplexed bead assay (MBA). Data were analyzed by multivariable logistic regression with both anti-LID-1 antibody positivity and history of HD as outcomes in separate models. Exposures in the analysis included history of parasites (both antibody results and self-reports) and several pertinent socio-demographics like area of residence (i.e. urban vs. rural).

**Results:** Of 1,311 enrollees, 72 (5.5%) reported a prior history of HD, 94 (7.2%) tested positive for anti-LID-1, 836 (63.8%) reported having one or more parasitic diseases in the past, 153 (11.7%) tested positive for antibodies to schistosoma egg antigen (SEA), and 69 (5.3%) for antibodies to the *Strongyloides stercoralis* antigen NIE. There was an association between rural residence and history of HD (aOR, 1.97, CI: 1.14 – 3.38). Rural residence and anti-LID-1 also showed a positive assocation (aOR 1.79, CI: 1.07-3.38). While not statistically significant, there was a positive association between anti-LID-1 and NIE antibodies (aOR 1.57, CI: 0.69-3.57), and a negative association between anti-LID-1 and SEA antibodies (aOR 0.79, CI: 0.38 - 1.61).

**Conclusion:** This study utilized a novel methodology (multiplex bead assay) to simultaneously measure seroreactivity among various pathogens. While we did not find that a history of HD and anti-LID-1 positivity were associated with seropositivity to NIE or SEA, our study found a high burden of several neglected tropical diseases (NTDs). There was also a strong association with rural residence and LID-1 positivity. This warrants further investigation into the prevalence and epidemiology of *M. leprae* infection, as well as potential spatial and environmental risk factors.

## INTRODUCTION

Hansen’s disease (HD), also commonly known as leprosy, is an ancient disease of humans [1] and is caused by the bacteria *Mycobacterium leprae* (and less commonly *Mycobacterium lepromatosis*). Each year there are approximately 200,000 new cases worldwide[2], with Brazil having the 2^nd^ highest incidence rate [3], despite significantly improved multi-drug treatment [4]. In 2023 Brazil had 22,773 annual new cases [5]. Brazil has had the highest incidence rate of Amazonian countries[6] and cases are disproportionally found in the Northeast within the country [7]. The exact mode of transmission is not fully understood; however, the most widely accepted theory is person-to-person through nasal secretions. There are also recent studies suggesting influence from environmental reservoirs [2, 6, 8, 9]. The epidemiologic association of high transmission in certain areas of Brazil is also not fully understood and may be due to specific unique risk factors present in these areas. Risk factors that may aggravate or facilitate transmission include poverty, undernutrition, water and sanitation practices, armadillo contact, and other environmental exposure [2, 7, 8, 10–12]. Another intriguing association, which we wanted to investigate here, is the possible increased susceptibility to HD due to pre-existing helminth infection.

It is not uncommon for populations in low-income settings to be burdened by multiple chronic infectious diseases simultaneously [13]. Helminth infections affect up to a quarter of the world’s population and primarily affect impoverished communities globally [14]. Schistosomiasis and intestinal helminths elicit a strong systemic T-helper cell (Th)-2 response which up-regulates regulatory T-cell activity, leading to a weakened pro-inflammatory Th1 response [15]. Since host control of *M. leprae* relies on a robust Th1 response, this may be a mechanism of risk for the development of HD. One study previously demonstrated that prevalence of multibacillary HD was much higher among patients with soil-transmitted helminth infections than those with no helminth infections[15]. In this situation, Th2 cytokines were more prominent in the co-infected individuals. Studies have demonstrated an association between HD and soil-transmitted helminths, in Brazil [16], Indonesia [17] and in West Africa [18]. In 2017 a study in Brazil demonstrated that evidence of a spatial overlap between HD and schistosomiasis[19]. These associations led us to investigate associations between HD and two helminths using serology.

To better understand the epidemiology of HD and select parasitic infections in eastern Minas Gerais, we utilized a multiplex bead assay to measure seroreactivity to *M. leprae*, *Schistosoma mansoni*, and *Strongyloides stercoralis*. *M. leprae* serological assays, including anti-LID-1 (IDRI Diagnostic protein) and anti-PGL1 (phenoglycolipid-1) can predict which individuals are at higher risk for developing HD, and presumably represent latent infection in those without symptoms [20, 21]. While the sensitivity and specificity of these tools for latent infection has not been determined, they can give us an epiodemiologic snapshot of HD and explore associations between seroreactivity and other factors, something that has not been fully investigated in the literature. This study aimed to determine the associations between concomitant *Schistosoma mansoni* and *Strongyloides stercoralis* antibody positivity with anti-LID-1 positivity to better understand the risks of *M. leprae* infection.

## METHODS

### Ethics Statement

This study was approved by the institutional review boards of Emory University and Universidade Federal de Juiz de Fora. All participants provided written informed consent with parents providing consent for their children. In addition, Children ages 7 -17 years old provided assent as per IRB guidelines. CDC staff were determined to be non-engaged in human subjects research as they did not interact with study participants or have access to identifying information.

### Study design and population

The study took place in four municipalities of eastern Minas Gerais, Brazil, in and near the city of Governador Valadares (Governador Valadares [GV], Inhapim, Teófilo Otoni, and Mantena). Areas were chosen due to relatively high rates of reported HD, with some areas considered hyperendemic (incidence rate of >40 / 100,000 annually)[8, 22, 23]. We partnered with local family health centers of each neighborhood or municipality to assist with recruitment of community members. Eligible participants included adults and children ages 3 years and older currently living in the communities. To have a representative sample for each neighborhood, households were chosen randomly across the neighborhoods with the assistance of the community health worker, ensuring geographic separation ( i.e. no contiguous households and on average 1-3 households per block). Invitiations were given for one individual per household to attend the recruitment and enrollment day later that week. While the goal was only one individual per household, rarely, more than one individual per household was enrolled.

### Data collection

Enrollment occurred from September 2021 to December 2022 with study activities conducted at local family health centers. A questionnaire was administered and included questions on demographics (i.e. sex, age, race, residence, monthly salary with minimum wage defined as 1,320 Reais per month [24]), household factors, risk factors for HD and self-reported history of the following infections: roundworm, schistosomiasis, hookworm, tapeworm, pinworm, giardia, amoebiasis, and HD. In addition, individuals were asked if they had a scar from the BCG (bacille Calmette-Guérin) vaccine, is given to household contacts (HHC) of individuals with HD because it can reduce the risk of the disease for HHC [25].

A fingerstick blood draw was taken for a dried blood spot which was transported to the Centers for Disease Control and Prevention in Atlanta, GA. Serum was eluted from the dried blood spots [26–31] and tested for antibodies to LID-1 from *M. leprae*, a pan-*Schistosoma* soluble egg antigen (SEA), and NIE from *Strongyloides stercoralis.* Using a multiplex bead serological assay (MBA) SEA and NIE were coupled to MagPlex beads (Luminex Corp, Austin, TX) as previously described [27, 28, 30, 31]. LID-1 was coupled with the same procedure using 15 µg of antigen per 12.5 × 10^6^ beads in PBS pH7.2. Seropositivity cutoffs for LID-1 and SEA were determined using the mean plus three standard deviations of readings from 85 individuals from a non-endemic area, while the cutoff for NIE was generated using receiver operator characteristic (ROC) curve analysis of 94 sera from non-endemic individuals and 53 from individuals whose stool was positive for *S. stercoralis* at the time of sera collection [32].

### Statistical analysis

All data were maintained on the data management platform RedCAP [33] and analyses were carried out in R4.3.0 [34] using packages: ‘readxl’, ‘janitor’, ‘tidyverse’, ‘AICcmodavg’, ‘gmodels’, and ‘stargazer’. Descriptive statistics were performed using frequency and measures of central tendency as appropriate. Univariate analyses with either logistic regression for continuous exposure variable(s) or Chi-square test for categorical variable(s) were conducted to determine associations with anti-LID-1 seropositivity or history of HD. For the purpose of further analyses, all categorical variables in the questionnaire were transformed to binary indicators, as follows: Sex as ‘female’ or ‘male’ (no one identified as non-binary), Race as ‘non-white’ (black, mixed, indigenous, Asian descent, other) or ‘white’, Residency as ‘rural’ or ‘urban’, Salary as ‘<1 minimum’ (1 x the minimum wage is a standard measure of monthly salary) or ‘>1 minimum’. Age was used as both a continuous variable and as a binary variable to create categories of participants less than the age of 15 (regarded as a minor) and all others over the age of 15 (regarded as an adult). Sample size calculations done apriori used an estimated odds ratio of 3 for the association of anti-LID-1 and anti-SEA (based on prior reports of associations between HD and helminth infection of 4 or more [15, 16]), and an estimated 10% seropositivity for *schistosoma* infection. The sample size calculation required to attain a power of 0.8 and significance of 0.5 was 45 individuals positive for anti-LID1 and 446 negative.

The crude forms of the models started with the following variables: ‘age’, ‘sex’, ‘race’, ‘residence’, ‘monthly salary’, ‘bcg scar’, ‘past parasitic infection’, self-reported parasites (‘Roundworm’, ‘Schistosomiasis’, ‘Hookworm’, ‘Tapeworm’, ‘Pinworm’, ‘Giardia’, ‘Amoeba’, and ‘Other’), ‘Strongyloides antibody (Anti-NIE)’, and ‘Schistosomiasis antibody (Anti-SEA)’. The variables ‘Reported History of Hansen’s Disease’ and ‘Antigen LID-1 antibody’ operated as the outcomes of interest. We reached our final models through backwards selection by dropping 1) ‘age’, ‘sex’, ‘monthly salary’, ‘past parasitic infection’, and ‘bcg scar’ for the Pos Anti-LID-1 model and 2) ‘race’, ‘monthly salary’, ‘past parasitic infection’, and ‘bcg scar’ for the Reported History of HD model.

Multivariate logistic regression models were built to explore associations between anti-LID-1 positivity and helminth serology (anti-SEA positivity and anti-NIE positivity) . We also built separate models with history of HD as the outcome instead of anti-LID-1 positivity. We then incorporated the following other measures of infectious parasites in three separate models: 1) individual infectious parasite that were self-reported from the questionnaire; 2) all individual infectious parasites into categories as follows: self-reported schistosomiasis, self-reported non-schistosomiasis helminths (comprising roundworm, hookworm, tapeworm, pinworm), and self-reported protozoa (comprising giardia, amoeba, and other); and 3) model that included only ‘yes’ or ‘no’ to whether someone had any history of parasitic disease. Each of these models incorporated two different outcomes of interest: 1) a participant resulting ‘positive’ for anti-LID-1 antibodies and 2) a participant having had a history of HD. Model diagnostics with backwards selection and Akaike information criterion (AIC) were used to determine which variables were used for the final models.

## RESULTS

### Demographics

A total of 1,315 participants were enrolled with 1,311 having sufficient data to include in the study. Four participants were removed for either having no results for the MBA assay or having not completed the surveys. The median age was 52 (range 4 to 92). 64.1% of participants were female (840/1311), 54.2% (710/1311) listed their race as ‘mixed’ , 68.3% (895/1311) had received BCG vaccination, 83.1% (1,089/1311) lived in an urban setting, and 24.3% (450/1311) made less than minimum wage for Brazil.

**Table 1** describes the descriptive statistics split into anti-LID-1 negative and anti-LID-1 positive. The majority of individuals were female and this did not differ between LID-1 groups (X^2^ = 0.38). Both antibody groups had similar results of one third of the participants being below minimum wage (X^2^ = 0.08). There were similar proportions from the LID-1 positive (83%) and negative (80.1%) who listed a non-white race (X^2^ = 0.42). Both LID-1 positive (68.1%) and LID-1 negative (68.3%) were relatively similar in their proportions of showing evidence of a BCG vaccination (X^2^ = 0.48). A larger percentage of the LID-1 positive group had a rural residence (26.6%) compared to their LID-1 negative counterparts (16%) (X^2^ = 6.94, p < 0.01).

**Table 1.**
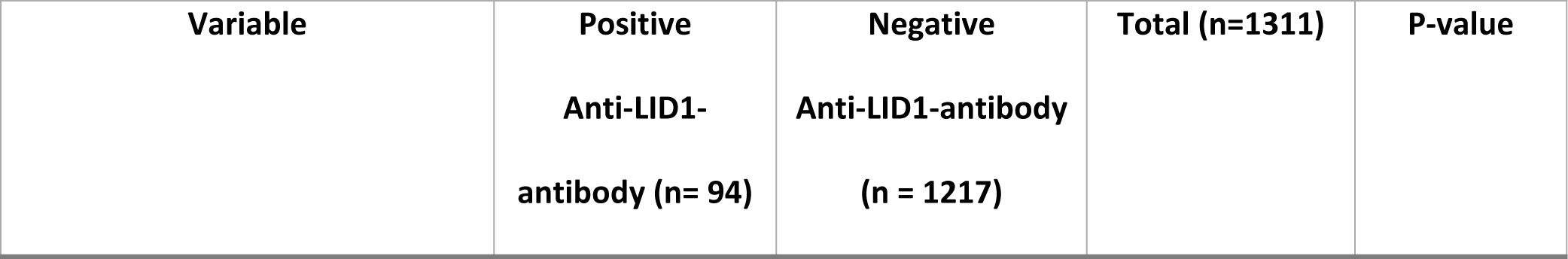
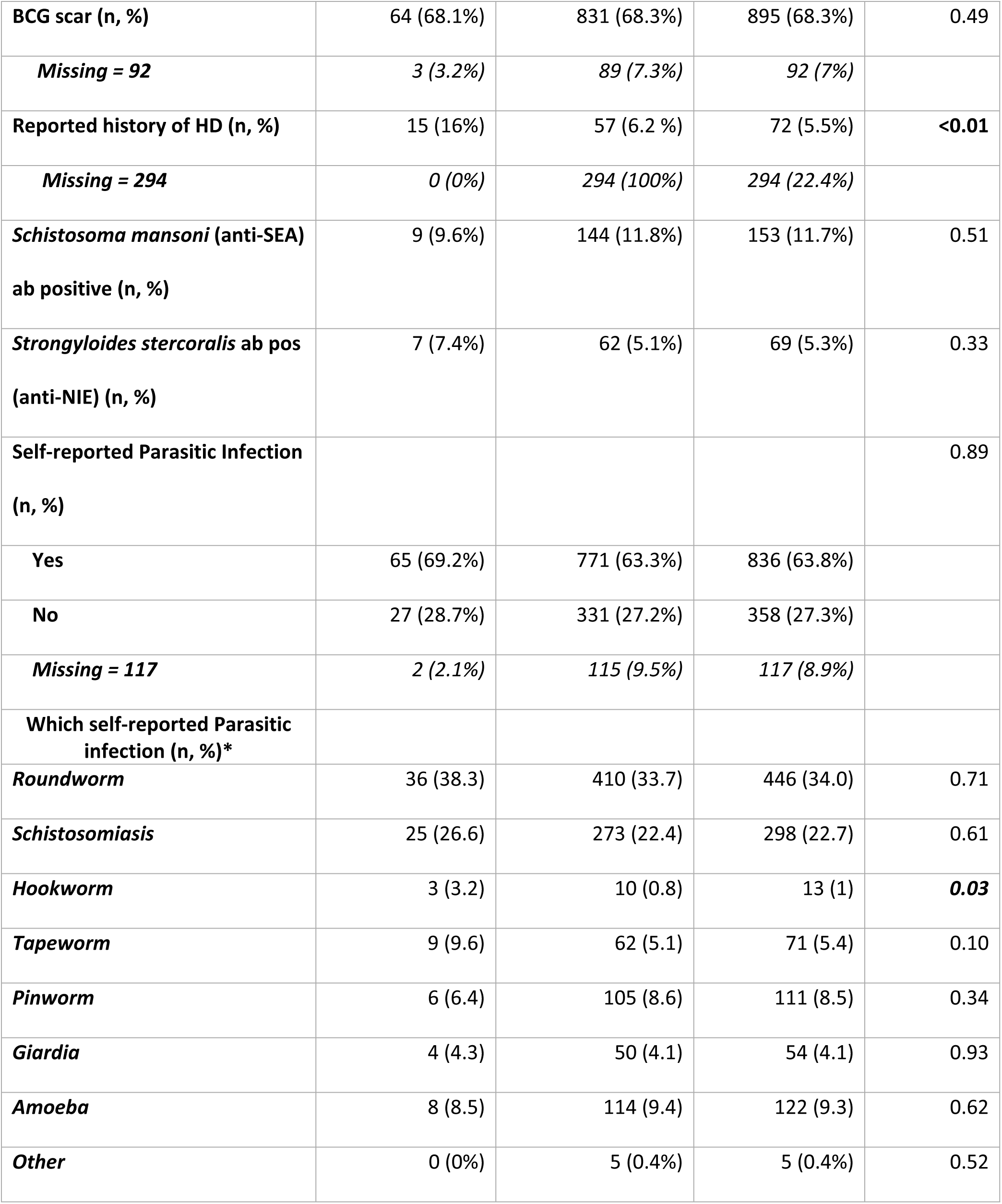
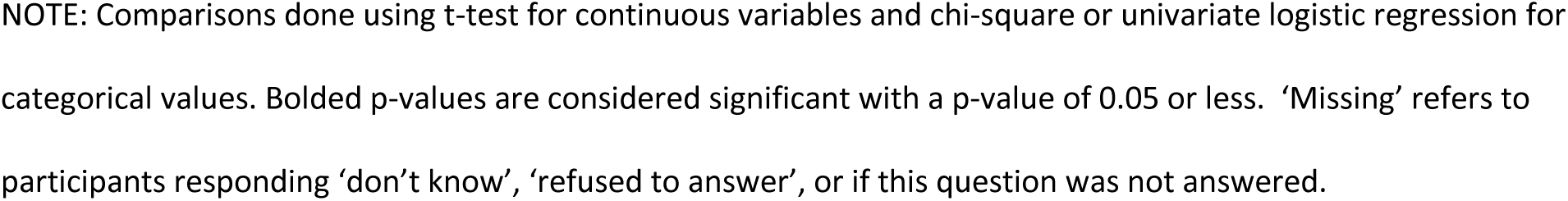
Demographic data comparing those who tested positive for anti-LID-1 antibody and those who tested negative.

| Variable | Positive<br>Anti-LID1-antibody (n= 94) | Negative<br>Anti-LID1-antibody<br>(n = 1217) | Total (n=1311) | P-value |
| --- | --- | --- | --- | --- |
| BCG scar (n, %) | 64 (68.1%) | 831 (68.3%) | 895 (68.3%) | 0.49 |
| *Missing = 92* | 3 (3.2%) | 89 (7.3%) | 92 (7%) |  |
| Reported history of HD (n, %) | 15 (16%) | 57 (6.2 %) | 72 (5.5%) | **<0.01** |
| *Missing = 294* | 0 (0%) | 294 (100%) | 294 (22.4%) |  |
| *Schistosoma mansoni* (anti-SEA)<br>ab positive (n, %) | 9 (9.6%) | 144 (11.8%) | 153 (11.7%) | 0.51 |
| *Strongyloides stercoralis* ab pos<br>(anti-NIE) (n, %) | 7 (7.4%) | 62 (5.1%) | 69 (5.3%) | 0.33 |
| Self-reported Parasitic Infection<br>(n, %) |  |  |  | 0.89 |
| Yes | 65 (69.2%) | 771 (63.3%) | 836 (63.8%) |  |
| No | 27 (28.7%) | 331 (27.2%) | 358 (27.3%) |  |
| *Missing = 117* | 2 (2.1%) | 115 (9.5%) | 117 (8.9%) |  |
| Which self-reported Parasitic<br>infection (n, %)* |  |  |  |  |
| *Roundworm* | 36 (38.3) | 410 (33.7) | 446 (34.0) | 0.71 |
| *Schistosomiasis* | 25 (26.6) | 273 (22.4) | 298 (22.7) | 0.61 |
| *Hookworm* | 3 (3.2) | 10 (0.8) | 13 (1) | **0.03** |
| *Tapeworm* | 9 (9.6) | 62 (5.1) | 71 (5.4) | 0.10 |
| *Pinworm* | 6 (6.4) | 105 (8.6) | 111 (8.5) | 0.34 |
| *Giardia* | 4 (4.3) | 50 (4.1) | 54 (4.1) | 0.93 |
| *Amoeba* | 8 (8.5) | 114 (9.4) | 122 (9.3) | 0.62 |
| *Other* | 0 (0%) | 5 (0.4%) | 5 (0.4%) | 0.52 |

NOTE: Comparisons done using t-test for continuous variables and chi-square or univariate logistic regression for categorical values. Bolded p-values are considered significant with a p-value of 0.05 or less. 'Missing' refers to participants responding 'don't know', 'refused to answer', or if this question was not answered.

### Infection history / status

**Table 2** provides the descriptive statistics of infectious disease data. Ninety-four individuals (7.2%) tested positive for anti-LID-1 antibody. The total number of individuals with a prior history of HD was 72 (5.5%), although this result was missing in 294 individuals since the question was introduced later in the study. Of these individuals reporting a history of HD, 15 (20.8%) were anti-LID-1 positive. For *S. mansoni* reactivity, 153 individuals (11.7%) tested positive, of which 9 individuals (9.6%) were anti-LID-1 positive. The association did not differ based on anti-LID-1 positivity (X^2^= 0.43, p = 0.51). Likewise, a total of 69 (5.3%) tested positive for *S. stercoralis* IgG, with 7 (7.4%) of those individuals also reacting seropositive for anti-LID-1, but the association also did not differ by anti-LID-1 positivity (X^2^= 0.97; p = 0.33).

**Table 2.** Infectious disease data comparing those who tested positive for anti-LID-1 antibody and those who tested negative.

| Variable | Positive<br>Anti-LID1-antibody (n= 94) | Negative<br>Anti-LID1-antibody<br>(n = 1217) | Total (n=1311) | P-value |
| --- | --- | --- | --- | --- |
| BCG scar (n, %) | 64 (68.1%) | 831 (68.3%) | 895 (68.3%) | 0.49 |
| *Missing = 92* | 3 (3.2%) | 89 (7.3%) | 92 (7%) |  |
| Reported history of HD (n, %) | 15 (16%) | 57 (6.2 %) | 72 (5.5%) | **<0.01** |
| *Missing = 294* | 0 (0%) | 294 (100%) | 294 (22.4%) |  |
| *Schistosoma mansoni* (anti-SEA)<br>ab positive (n, %) | 9 (9.6%) | 144 (11.8%) | 153 (11.7%) | 0.51 |
| *Strongyloides stercoralis* ab pos<br>(anti-NIE) (n, %) | 7 (7.4%) | 62 (5.1%) | 69 (5.3%) | 0.33 |
| Self-reported Parasitic Infection<br>(n, %) |  |  |  | 0.89 |
| Yes | 65 (69.2%) | 771 (63.3%) | 836 (63.8%) |  |
| No | 27 (28.7%) | 331 (27.2%) | 358 (27.3%) |  |
| *Missing = 117* | 2 (2.1%) | 115 (9.5%) | 117 (8.9%) |  |

| Which self-reported Parasitic infection (n, %)* |  |  |  |  |
| --- | --- | --- | --- | --- |
| **Roundworm** | 36 (38.3) | 410 (33.7) | 446 (34.0) | 0.71 |
| **Schistosomiasis** | 25 (26.6) | 273 (22.4) | 298 (22.7) | 0.61 |
| **Hookworm** | 3 (3.2) | 10 (0.8) | 13 (1) | **0.03** |
| **Tapeworm** | 9 (9.6) | 62 (5.1) | 71 (5.4) | 0.10 |
| **Pinworm** | 6 (6.4) | 105 (8.6) | 111 (8.5) | 0.34 |
| **Giardia** | 4 (4.3) | 50 (4.1) | 54 (4.1) | 0.93 |
| **Amoeba** | 8 (8.5) | 114 (9.4) | 122 (9.3) | 0.62 |
| **Other** | 0 (0%) | 5 (0.4%) | 5 (0.4%) | 0.52 |

\*Self-reported refers to participants who answered 'Yes' or 'No' to the following question – "Have you ever been diagnosed with a parasitic infection?". If a participant selected 'Yes' they were then prompted with a list of possible parasites and to indicate which of the following they had been infected by and/or provide an option for 'Other' if a parasite was not listed. 'Missing' refers to participants responding 'don't know', 'refused to answer', or if this question was not answered
NOTE: Comparisons done using t-test for continuous variables and chi-square or univariate logistic regression for categorical values. Bolded p-values are considered significant with a p-value of 0.05 or less.

In terms of self-reported history, almost two-thirds, almost 2/3 of individuals reported a prior parasitic infection (63.8%), with roundworm (34%) and schistosomiasis (22.7%). being the most common. Interestingly, only 47 (15.8%) of those reporting a history of schistosomiasis tested positive for anti-SEA antibodies, possibly signifying a more remote history or clearance of antibodies due to treatment. There was a positive association between self-reported hookworm and LID-1 seropositivity (X^2^= 4.37; p = 0.03).

For the option ‘other diseases’ five people responded, which included the following answers: 2 for leishmaniasis, 1 for toxoplasma, 1 for urinary infection, and 1 response that was unknown. In this study, 117 participants were excluded from analysis of data related to reported past parasitic infection because they marked ‘don’t know’(97), ‘refuse to answer’ (15), or had not answered the question at all (5). **Table 3** shows all the responses for the multivariate logistic models for both outcomes of interest.

**Table 3:** Multivariate Logistic Regression with POSITIVE ANTI-LID1 and HISTORY OF HANSEN’S DIAGNOSIS as outcomes (serological results and categorized parasites).

|  | Pos Anti-LID-1 | History of HD |
| --- | --- | --- |
|  | aOR (95% CI) | aOR (95% CI) |
| *S. mansoni* antibody positive (anti-SEA) | 0.79 (0.38, 1.61) | 1.26 (0.63, 2.50) |
| *S. sterocralis* antibody positive (anti-NIE) | 1.57 (0.69, 3.57) | 0.40 (0.09, 1.70) |
| Self-reported schistosomiasis | 1.02 (0.61, 1.70) | 0.88 (0.50, 1.53) |
| Self reported helminth infection (other than *S. mansoni*) | 1.15 (0.74, 1.78) | **1.77 (1.08, 2.90)** |
| Self-reported protozoa | 0.76 (0.37, 1.57) | 0.75 (0.35, 1.62) |
| Race (non-white) | 1.20 (0.68, 2.10) |  |
| Adult age (15 or older) |  | **7.80 (1.05, 57.64)** |
| Sex as female |  | 0.64 (0.39, 1.051) |
| Rural Residence | **1.79 (1.07, 2.99)** | **1.97 (1.14, 3.38)** |

Note: Adjusted odds ratio listed with confidence intervals (in parentheses) of exposures involving positive serological results of helminth infections, categorized self-reported past or present infections, and best fit demographic variables. Categorization of parasites was as follows: self-reported *S.mansoni* infection, self-reported Helminth Infections other than *S.mansoni* (Roundworm, Hookworm, Pinworm, Tapeworm), and self-reported Protozoa Infections (Giardia, Amoeba, Other). \* p<0.05

### Multivariate analysis

Final logistic regression models are shown in **Table 3** involve the categorized form of the self-reported parasites. Two other models for each outcome were run and are displayed in supplementary **Tables S1 and S2**, comprising the forms of each indivualized parasite and all parasites summized into the singular variable ‘parasitic infection’. The final risk factors for each model type are those not dropped during backwards selection and the exposures kept a priori (‘Strongyloides antibody [Anti-NIE]’, ‘Schistosomiasis antibody [Anti-SEA]’, and self-reported parasites). Since the anti-SEA variable and self-report parasite variable for schistosomiasis had the potential for multicollinerarity, we ran additional permutations of the selected models. This included the original form (with both thenti-SEA antibody and the self-reported schistosomiasis), a version of the model with only the anti-SEA antibody, and a version with only the schistosomiasis self-report variable. There was little to no effective change, so we determined to there was not concern for collinearity with the original selected model. **Tables S3 and S4** describe these permutations and can be found in the Supplementary Section.

Anti-LID1 positivity was positively associated with anti-NIE positivity (aOR 1.57; 95% CI:0.69 - 3.57), although this did not reach statistical significance. Positive anti-SEA antibody results showed a negative association (aOR 0.79, 95% CI: 0.38 - 1.61) with anti-LID-1 positivity, but again not significant. Self-reported parasitic infections (categorized by type) were not associated with anti-LID-1 positivity. Finally, participants with positive anti-LID-1 (aOR 1.79, 95% CI: 1.07, 2.99) were more likely to live in a rural locale than urban based participants (**Table 3**).

For the outcome History of Hansen’s Disease there was a positive association (aOR 1.26, 95% CI: 0.63 - 2.50) of having *S. mansoni* antibodies and a negative association (aOR 0.40, 95% CI: 0.09 - 1.70) of *Strongyloides sp.* antibodies, although neither statistically significant (Table 3). Self-report of non-*Schistosoma* helminth infection was associated with prior HD (aOR 1.77, 95% CI: 1.08 - 2.90), as was older age (15 and greater) (aOR: 7.80, 95% CI 1.06 - 57.6). Participants with prior history of Hansen’s Disease (aOR 1.97, 95% CI: 1.14 - 3.38) were more likely to reside in rural areas than urban settings (**Table 3**).

## DISCUSSION

Our study showed a high burden of HD in eastern Minas Gerais with more than 5% of a random sample having had prior Hansen’s disease and almost 8% with seroreactivity to *Mycobacterium leprae* through anti-LID-1 antibody. Seroreactivity *to S. mansoni* and *S. stercoralis* was observed in 5-12% of participants, and almost two-thirds of participants reported a past parasitic infection (both NTDs and others). Studies using anti-LID-1 serology for assessing community prevalence are not common, and none have used a multiplexed beaded assay (MBA) to measure serology. One other study in Brazil found that over 20% of the study population tested positive for anti-LID-1 [20], which is more than twice as high compared to our findings of 7%. However, our study is more in line with a study on household contacts in Goais, Brazil, which found 7% seropositivity [35]. While age did not play a role in our analysis to the LID-1 outcome, one study interestingly found that youngest age groups (under 30 years old) had statistically significant higher *M. leprae* seropositivity (as measured by anti-PGL1) than older groups [36]. We did, however, find a strong association with age and history of HD, consistent with prior literiature of increased HD in older populations, although we did not know the exact age of acquisition of HD [37, 38]. Our findings suggest that acquisition of infection is not necessarily age related, but that progression to disease is.

Our MBA provided the ability to measure seropositivity of several pathogens at once, however, we did not find statistically significant associations with serology for helminths and either LID-1 or prior history of HD as hypothesized. Co-infections with helminths, particularly soil-transmitted infections and schistosomiasis, have been shown to have epidemiologic and immunologic associations with Hansen’s Disease in prior research[13, 15, 16].

*S.mansoni* is an endemic helminth infection in Minas Gerais [39], the only schistosoma species in Brazil, and was highly self-reported in our study (23%). In addition, 12% of individuals had a positive anti-SEA IgG, signifying a treated (resolved) or chronic *S. mansoni* infection. Having had a history of HD was positively associated with anti-SEA seroreactivity (aOR 1.26, 95% CI: 0.63-2.50), but did not reach statistical significance, making it difficult to make any definitive conclusions. Our prior work in this region demonstrated a strong association between active *S. mansoni* infection and HD (aOR 8.69, 95% CI: 1.50 – 50.51) when comparing cases to household contacts [16], therefore, continued investigation into potential mechanisms and temporal associations is warranted. Not knowing the time of onset of either infection in the current study (*M. leprae* or *S. mansoni*) may have masked potential associations.

*S. stercoralis*, a soil-transmitted helminth found mostly in tropical/subtropical zones with moist soil and poor sanitation [40], was the other directly measured helminth infection in this study. A positive serological result could be reflective of a past or current, since it is hard to diagnose on stool exam [41–43] and over 5% of study participants tested seropositive for this parasite. While not statistically significant, anti-NIE seroreactivity was positively associated with anti-LID-1 (aOR 1.57, 95% CI: 0.69-3.57), and deserves further study, especially with the associations in the literature between soil-transmitted helminths and HD.

To have a better idea of prior exposures in addition to the measured antibodies, we asked participants to report any prior diagnoses of parasitic infections. Interestingly, self-reported worm infections (excluding schistosomiasis) had a stasticially significant positive assocation with history of HD (aOR 1.77, 95% CI: 1.08 – 2.90), while hookworm alone had a statistically significant positive aossication with anti-LID-1 seropositivity (OR 3.97, 95% CI: 1.08-14.7; p = 0.03). However, with very small numbers, specifically only 13 participants reporting a history of hookworm infection and with 3 of those individuals being anti-LID-1 positive, this limits interpretability of this difference. Again, these prior research has shown a positive assocation between HD and soil-transmitted helminths [15, 17, 44], and both of these findings with self-reported disease deserve further study.

We did not find an association with the category self-reported history of protozoa to either outcome of interest. There are few studies related to co-infection between HD and protozoa, one being that *Toxoplasma gondii* could potentially increase the chances of a more severe complication to HD [45]. More direct measurement of intestinal protozoa at the time of HD diagnosis could better determine an association. Lastly, the BCG vaccine has been shown to reduce the likelihood of HD in household contacts [25]. However, the presence of a BCG scar, indicating vaccination, did not differ across anti-LID-1 positive or negative individuals, suggesting that past vaccination was not associated with reduced risk of *M. leprae* infection.

In terms of non-infectious variables, there was a consistent significant association across all the models demonstrating a positive relationship between rural residence and both anti-LID-1 positivity and past HD. This result could be reflective of the endemicity in the rural areas and/or another risk factor such as environmental exposures.

There were several limitations of this study. Firstly, this analysis is a cross-sectional study, making it relatively difficult to establish a causal relationship between the exposures and outcomes. The variables related to having a history of parasites as well as history of past Hansen’s Disease diagnosis involved recall bias due to the nature of self-reported data. Additionally, the history of Hansen’s Disease was missing nearly 300 responses, which may have impacted the power of the analysis for this outcome.

## Conclusion

This study utilized a novel multiplex bead assay to simultaneously measure seroreactivity to *M. leprae*, schistosomiasis, and strongyloidiasis in a community sample and to examine associations between the infections. While we did not find strong associations, the degree of overlap of infections and baseline seropositivity of three NTDs in this region are notable. The strong association with rural residence also warrants further investigation by geospatial techniques to identify spatial risks or environmental risk factors. These epidemiologic tools, thus, hold promise for wider use in other areas to understand the prevalence of *M. leprae* infection and potential other infections that impact disease epidemiology.

## Data Availability

All relevant data are within the manuscript and its Supporting Information files. But we do not want to publish these data if accepted. We will make available elsewhere

## Acknowledgements

We thank Diana Martin and Gretchen Cooley at the U.S. Centers for Disease Control and Prention, for their assistance with the serum testing on the multiplexed beaded assay. We also appreciate the community health workers and other healthcare providers who assisted with recruitment and use of their healthcare facilities for the study. Finally, the project could not have happened without the willing participation of the community members themselves.

## SUPPLEMENTARY TABLES AND FIGURES

**Supplementary Table 1:**
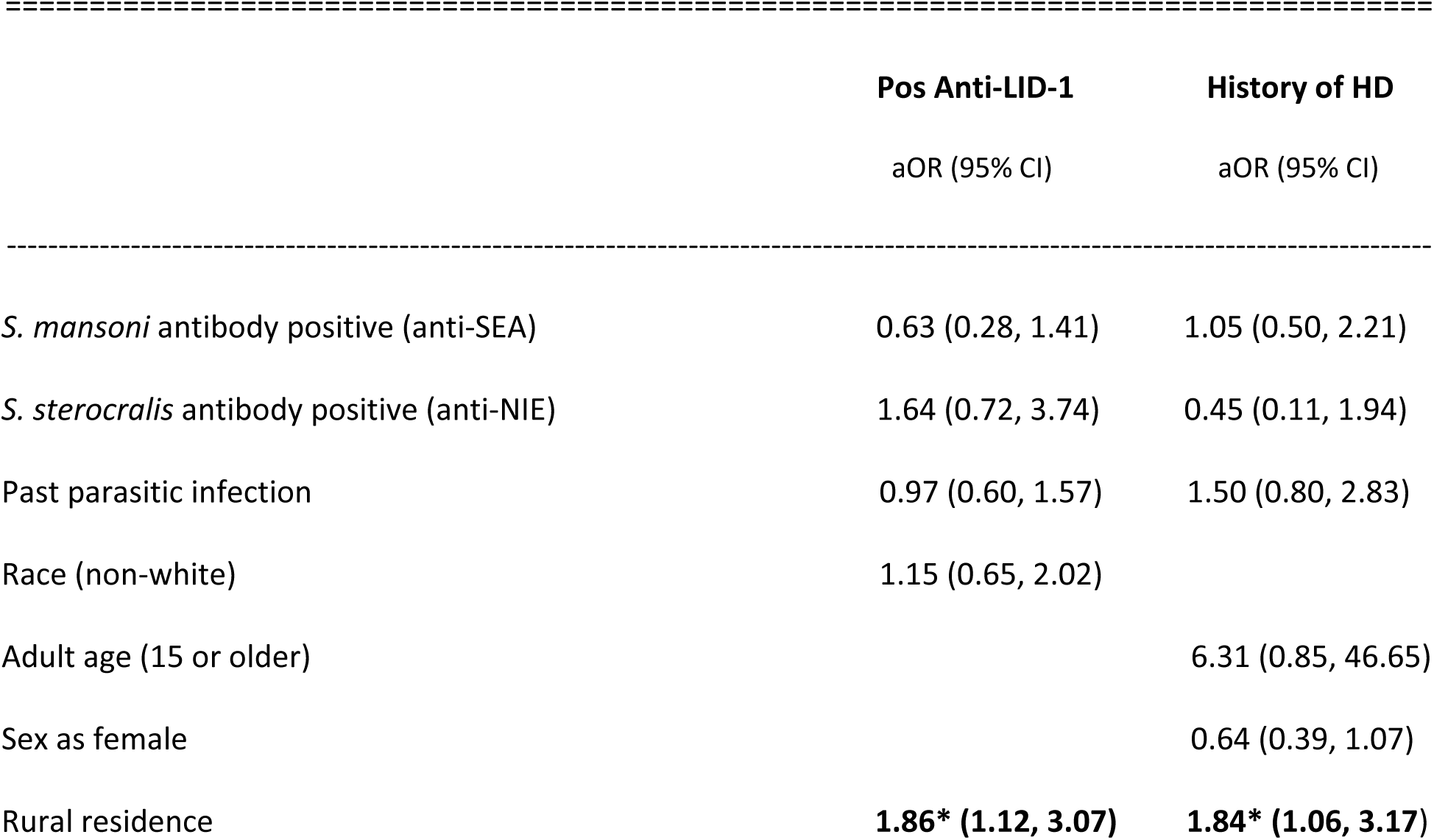
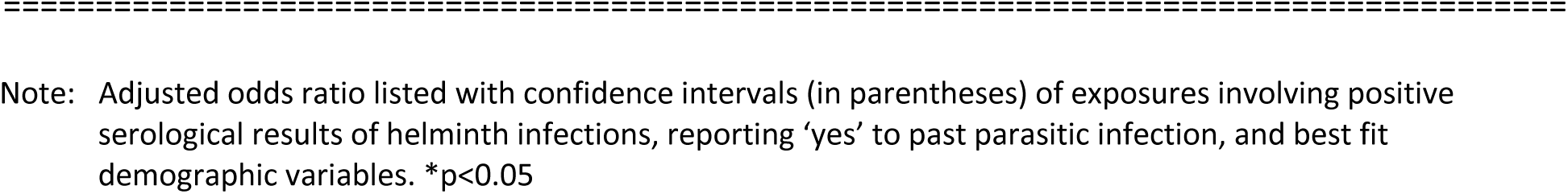
Multivariate Logistic Regression with <u>POSITIVE ANTI-LID1</u> and <u>HISTORY OF HANSEN’S DISEASE</u> against co-infection (serological results and *past parasitic infection*) and demographic risk factors.

|  | Pos Anti-LID-1 | History of HD |
| --- | --- | --- |
|  | aOR (95% CI) | aOR (95% CI) |
| *S. mansoni* antibody positive (anti-SEA) | 0.63 (0.28, 1.41) | 1.05 (0.50, 2.21) |
| *S. sterocralis* antibody positive (anti-NIE) | 1.64 (0.72, 3.74) | 0.45 (0.11, 1.94) |
| Past parasitic infection | 0.97 (0.60, 1.57) | 1.50 (0.80, 2.83) |
| Race (non-white) | 1.15 (0.65, 2.02) |  |
| Adult age (15 or older) |  | 6.31 (0.85, 46.65) |
| Sex as female |  | 0.64 (0.39, 1.07) |
| Rural residence | **1.86\* (1.12, 3.07)** | **1.84\* (1.06, 3.17)** |

Note: Adjusted odds ratio listed with confidence intervals (in parentheses) of exposures involving positive serological results of helminth infections, reporting 'yes' to past parasitic infection, and best fit demographic variables. \*p<0.05

**Supplementary Table 2:**
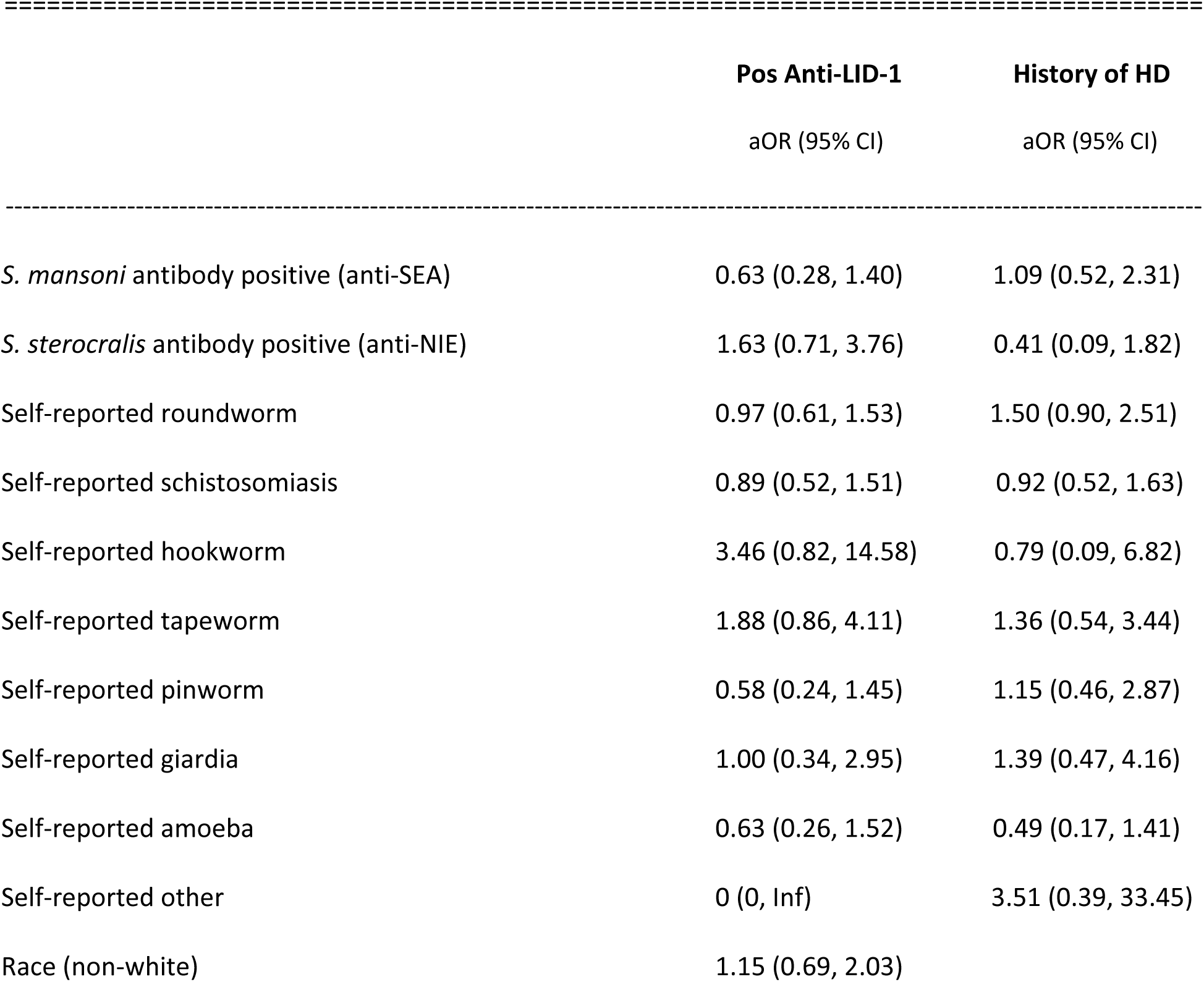
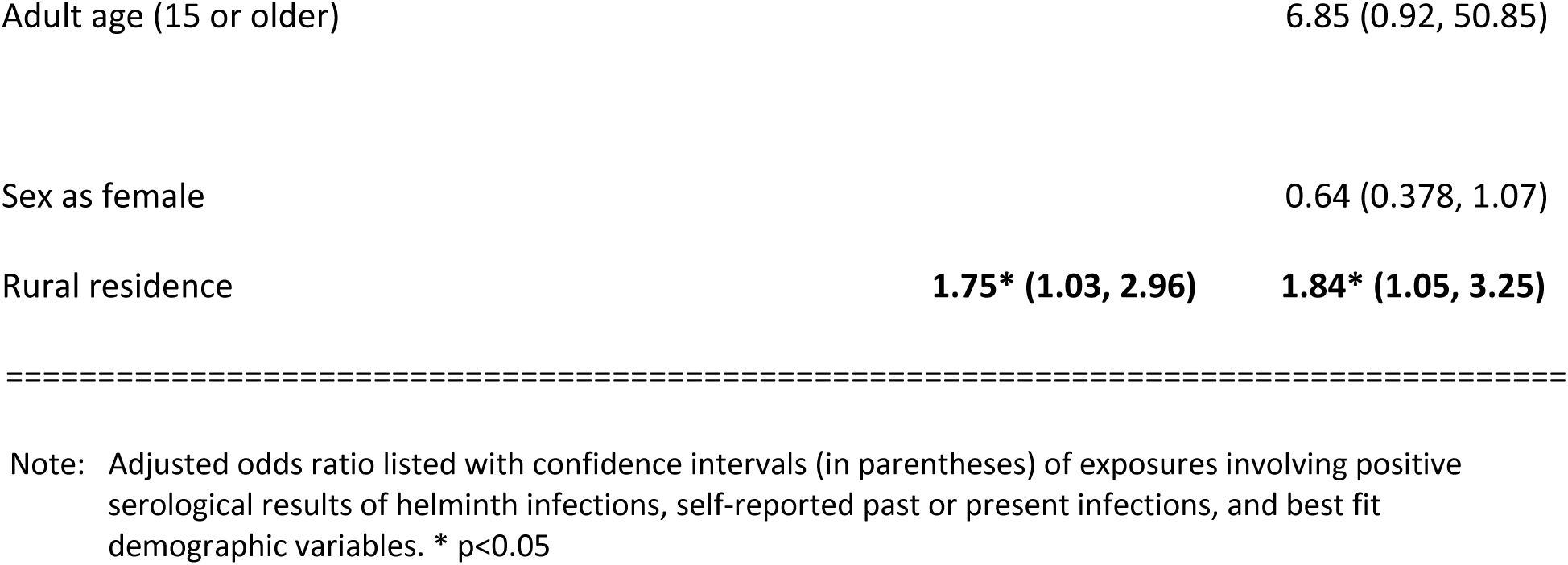
MLR with <u>POSITIVE ANTI-LID1</u> and <u>HISTORY OF HANSEN’S DISEASE</u> against co-infection (serological results and *all individual parasites*) and demographic risk factors.

**Supplementary Table 3:** Checking effects size change among different model permutations for the outcome of POSITIVE ANTI-LID1 (aOR)

| 455 |  | Both A & SR | A Only | SR Only |
| --- | --- | --- | --- | --- |
| 456 | ----- |  |  |  |
| 457 | *S. mansoni* antibody positive (anti-SEA) | 0.78 |  | 0.78 |
| 458 |  |  |  |  |
| 459 | *S. sterocralis* antibody positive (anti-NIE) | 1.57 | 1.54 | 1.57 |
| 460 |  |  |  |  |
| 461 | Self-reported schistosomiasis | 1.00 | 0.99 |  |
| 462 |  |  |  |  |
| 463 | Self reported helminth infection (other than *S. mansoni*) | 1.14 | 1.15 | 1.14 |
| 464 |  |  |  |  |
| 465 | Self-reported protozoa | 0.96 | 0.95 | 0.96 |
| 466 |  |  |  |  |
| 467 | Race (non-white) | 1.21 | 1.21 | 1.21 |
| 468 |  |  |  |  |
| 469 | Rural Residence | 1.81** | 1.79** | 1.81** |

Note: 'A' stands for antibody result for schistosomiasis. 'SR' is self report for schistomiasis. \*\*p<0.05.

**Supplementary Table 4:**
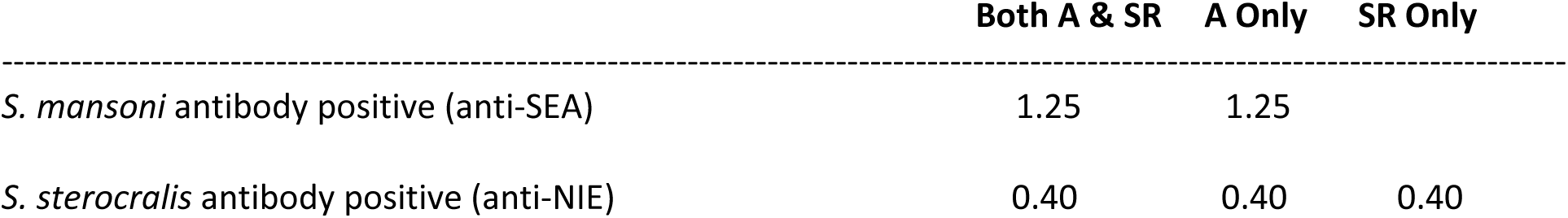
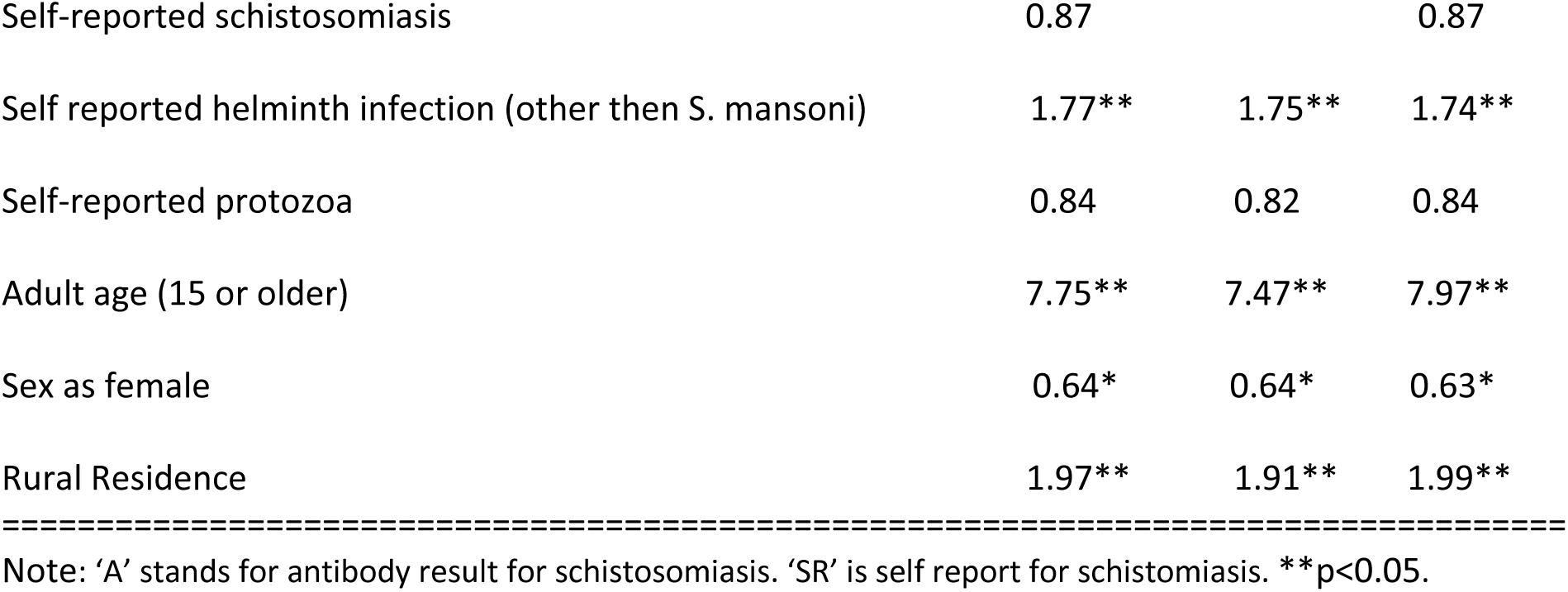
Checking effects size change among different model permutations for the outcome of HISTORY OF HANSEN’S DISEASE (aOR)

| 478 |  | Both A & SR | A Only | SR Only |
| --- | --- | --- | --- | --- |
| 479 | ----- |  |  |  |
| 480 | *S. mansoni* antibody positive (anti-SEA) | 1.25 | 1.25 |  |
| 481 |  |  |  |  |
| 482 | *S. sterocralis* antibody positive (anti-NIE) | 0.40 | 0.40 | 0.40 |

|  |  |  |  |
| --- | --- | --- | --- |
| Self-reported schistosomiasis | 0.87 |  | 0.87 |
| Self reported helminth infection (other than *S. mansoni*) | 1.77** | 1.75** | 1.74** |
| Self-reported protozoa | 0.84 | 0.82 | 0.84 |
| Adult age (15 or older) | 7.75** | 7.47** | 7.97** |
| Sex as female | 0.64* | 0.64* | 0.63* |
| Rural Residence | 1.97** | 1.91** | 1.99** |

Note: 'A' stands for antibody result for schistosomiasis. 'SR' is self report for schistomiasis. \*\*p<0.05.

